# COVID-19 Transmission Dynamics in India with Extended SEIR Model

**DOI:** 10.1101/2020.08.15.20175703

**Authors:** Brahmatheja Reddy Mali Reddy, Anuj Singh, Pradeep Srivastava

## Abstract

India is one of the most harshly affected countries due to COVID epidemic. Early implementation of lockdown protocols were useful to control certain parameters of transmission dynamics, but the numbers are consistently increasing in later months. India’s population is divided into different clusters on the basis of population density and population mobility, even varying resource availability and since the recent cases are coming from throughout the country, it allows us to model an overall average of the country. In this study, we try to prove the efficiency of using the SEIR epidemiological model for different rate study analysis for COVID epidemic in India. Along with it we derived newer components for better forecast of the pandemic in India. We found that there is a decrease in R_0_ value, but still the epidemic is not under control. The percentage of infected patients being admitted into ICU for critical care is around 9.986%, while the chances of recovery of critical patients being admitted to the ICU seem to be slim at 79.9% of the admitted being dead.

## Introduction

Originating in China during the end of 2019, COVID-19 is being called the world’s major problem. Everything from public health to the global economy is badly affected. The disease is caused by a novel virus called SARSCoV-2 (severe acute respiratory syndrome coronavirus 2). The first case was officially reported on 31st December 2019 and later after one month, it has declared as the world’s major health hazard by the World’s Health Organisation [1]. Till the date of August 13, the disease has spread and affected 213 countries or territories around the world [12]. During early 2020, China was the first country to record the COVID-19 infectious number up to 80,000 followed by France, Italy and other European countries [11]. They had their peak during March and April but somehow, they managed to lower down the number of active cases and the average of the new number of cases in a day dropped to less than 1000 (for the period of May, June and July)[12]. In the case of India, the numbers are initially lower (during the period of March April) because of lockdown implementation and inadequate testing facility in India. But in later months, India has recorded big numbers in COVID tally and because of higher population density, cities like Mumbai, Delhi, Chennai and Ahmedabad contributed most of the numbers in the country’s total[7,8]. The Indian population is divided among different clusters and therefore, there is a difference in a pandemic in different states. Recently, some of the states have seen flood issues (Assam and Bihar) which created a sudden disturbance in social distancing measures and ultimately there is an increase in the number of cases from those states[9].

For India, during the first phase of CoronaVirus Pandemic, the numbers were added by few major cities but now, a small number is added by every district in India and therefore, India has an average of more than 60,000 cases in the last 10 days (from August 03^rd^ to August 13^th^) whereas it was an average of less than 10,000 in earlier months [8]. Since the COVID has spread across the country, so it is better to model it through an epidemiological method. There are several studies or models in epidemiology, where they try to understand the behaviour of disease spread [3,4,12]. These existing models are a general epidemic model to get an idea of transmission dynamics of a pandemic, stages of the pandemic and what shape it would take with time. Some of the very popular epidemiology models are SIR, SIS and SEIR [5,6].

Pandemic and infection spread is always dependent on the type of disease. In the case of COVID-19, it is an airborne disease and the virus directly targets the lungs. Therefore, its spread rate is quite higher than other infectious diseases. Secondly, the infected person gets to know about the infection in about 10 days on an average (in the case of non-asymptotic infection). Symptoms take 7 to 8 days on an average to show up and till then the individual could have spread the infection to other populations. There are the two most dominating factors for exponential growth in the total number of new cases day by day. For our current paper, we tried to understand the pandemic behaviour in India on the basis of the SEIR model. Huo HF et. al. studied the dynamics of the sexually transmitted disease using SEIR epidemiological model [13]. There are four main elements in the SEIR model viz. Susceptible (S), Exposed (E), Infectious (I) and Recovered (R). Apart from its limitations in applying to real scenarios, it is mostly used to analyse the COVID-19 pandemic by countries across the world [14,15,16].

## Method

In the SEIR Model, the transition from one compartment to another depends on the rate, probability and population. The rate describes how long a change takes, the population is the group of individuals that this change applies to and probability is the possibility of transmission taking place for an individual. Infectious diseases tend to spread from one group of population to another. Thus it is necessary to gain insights on their rate of spread, what proportion of a population is susceptible, what proportion is infected, etc. A compartmental model helps us separate the population into several components. A basic SEIR model resembling the scale of COVID-19 transmission dynamics is divided into 4 population compartments:

- Susceptible (S) (Healthy population, vulnerable to be infected)
- Exposed (E)
- Infected (I)
- Recovered (R)

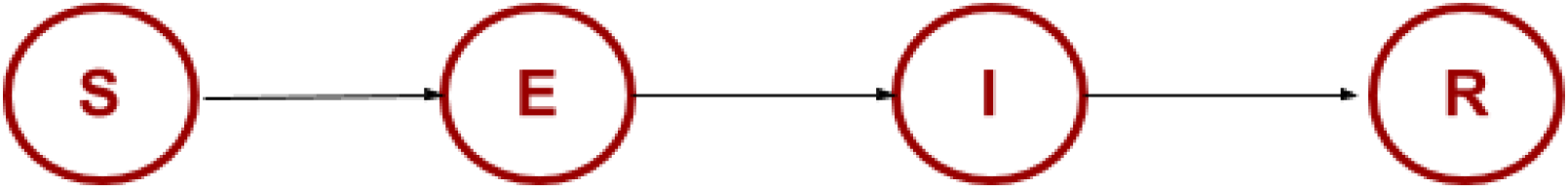

This is a SEIR model where there are four possible transitions between defined compartments.

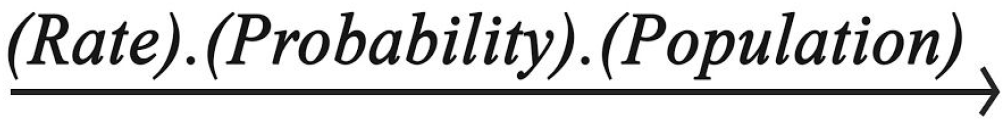

Here are all the parameters our model needs (that’s just all the variables in the equations plus the variables in the functions for R_0_(t) and Beds(t):

**Table 1:** List of all system variables defined in the Model.

| System Variables |  |
| --- | --- |
| $N$ | Total Population |
| $S(t)$ | Number of people Susceptible on day $t$ |
| $E(t)$ | Number of individuals Exposed on day $t$ |
| $I(t)$ | Number of Infected individuals on day $t$ |
| $R(t)$ | Recovery count on day $t$ |
| $D(t)$ | Number of deaths on day $t$ |

- **p(I→C):** the probability of going from infected to critical
- **p(C→D):** the probability of dying while critical

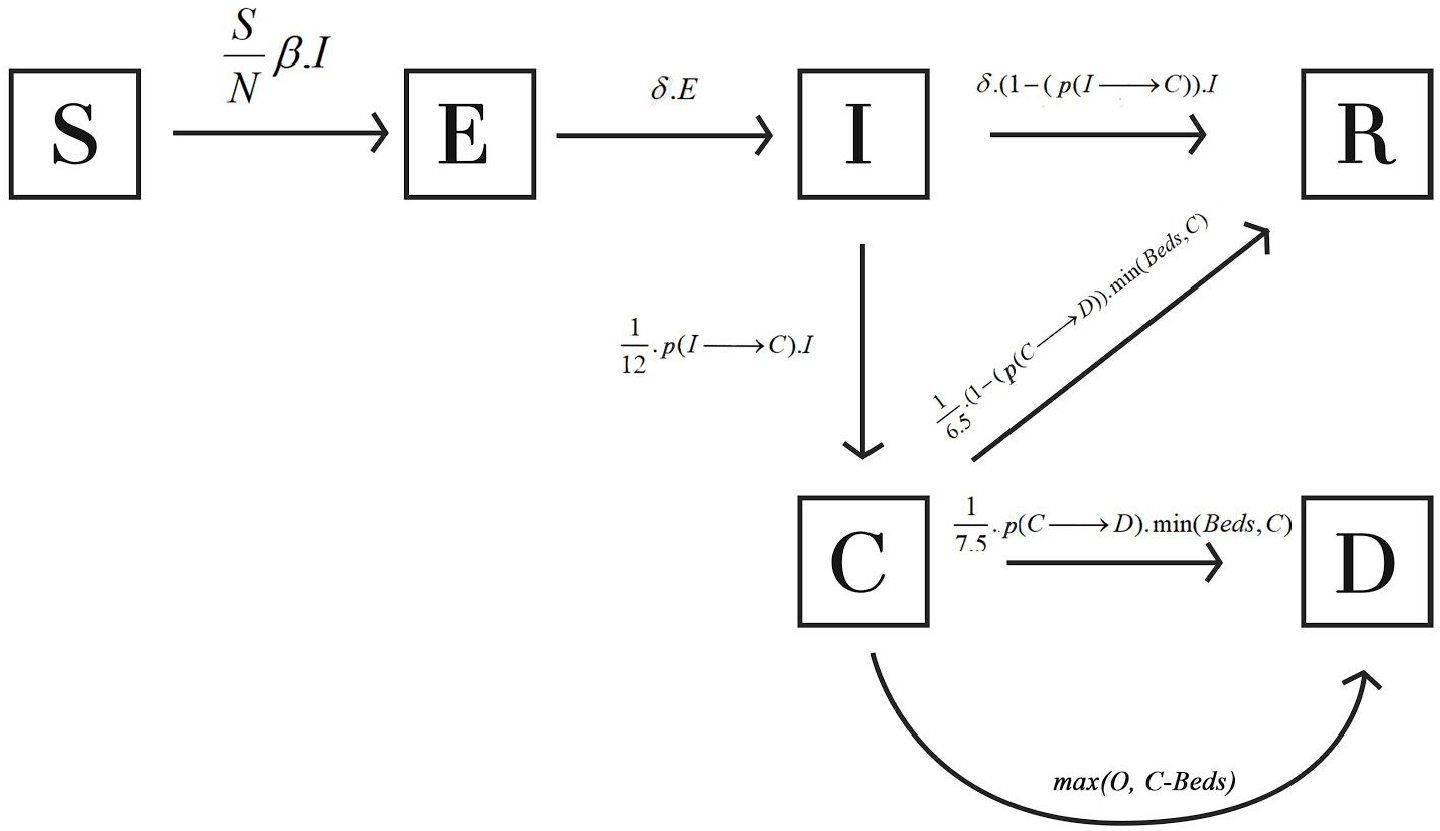

The following differential equations show the transitions from one compartment to other (mathematical expression for SEIR extended model):

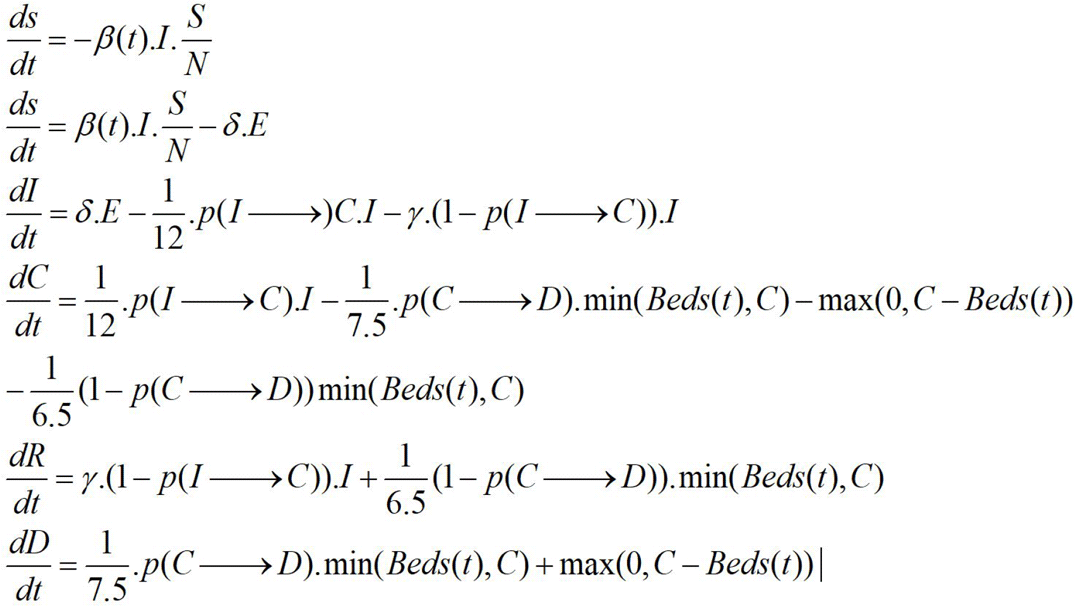

**Table 2:** List of all parameters that are considered in the Model.

| Parameters |  |
| --- | --- |
| $\beta$ | Expected number of people an infected person infects per day |
| $\gamma$ | The proportion of infected recoveries per day |
| $R_0$ | The total number of people an infected person infects |
| $\delta$ | Length of the incubation period |
| $\alpha$ | Fatality Rate |
| $\varrho$ | Rate at which infected people die. |
| $D$ | Number of days an infected individual has and can spread the disease |
| $R_{0start}$ | Parameter in $R_0(t)$ |
| $R_{0end}$ | Parameter in $R_0(t)$ |
| $x_0$ | Parameter in $R_0(t)$ |
| $k$ | Parameter in $R_0(t)$ |
| **$Beds(t)$** | Number of Beds at day $(t)$ |
| **$Beds_0(t)$** | Parameter in $R_0(t)$ |

- The *beds’* dataset has the amount of ICU beds *per 100k inhabitants* for many countries. [min of health]
- The *age groups* dataset has the number of people per age group for all countries. [census india]
- The *probabilities* table has probabilities containing I→C and C→D per age group. [min of health]
- *covid_data* is a huge table with the number of fatalities per region per day, from 22nd January 2020 onwards. [min of health]

## Deriving the Exposed-Compartment

In the case of COVID-19, the disease can have an incubation period before getting infected. The period when the infected individual has not spread the virus to other individuals is called the Exposed compartment in the SEIR epidemiological model. There is a transition sequence, as contact with Infected ones and then they are in the Exposed compartment. This is a transition shown in **figure 3**, which is the key to the SEIR model. The Susceptible population can come in contact with Infected ones and then they are in the Exposed compartment. This is a transition from Susceptible to Exposed i.e. S → E. The transition S → I will have almost the same probability because exposition can happen immediately and the total population remains the same. Mathematically, an infected person can spread the disease and every individual exposes new ones per day. In our case, every exposure becomes infected and the rate becomes ***δ***.

**Figure 3:**
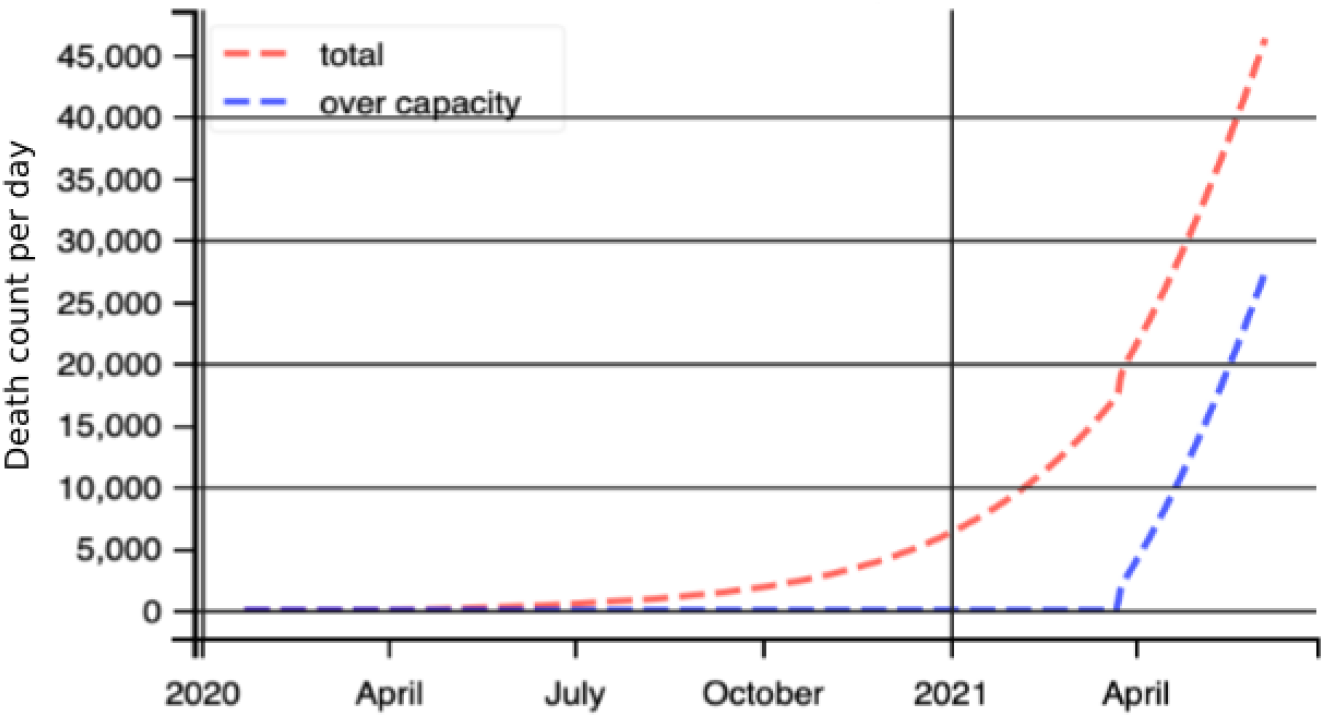
Deaths per day projection with time for India

## Deriving Dead Compartment

For coronavirus disease, a dead compartment is significant and crucial because of the non-availability of known treatment. There is quite a higher number of deaths in a day in India for the last few weeks. While adding a transition from Infectious to Death i.e. I → D, we define a new parameter ***ϱ ° ϱ*** can be defined as the rate of the deceased case (for example, when an infected individual takes five days to die, the ***ϱ*** will be 1/5). Apart from the rate of the deceased case, there would be no change in the recovery rate. The death rate ***α*** will be 1 – recovery rate. In other words,

Recovery rate can be referred to as the probability of transition from Infectious to Recovered i.e. I → R. Similarly, the Death rate can be referred to the probability of transition I → D. The sum of both the probability is always 1.

## Time-Dependent R_0_

R tends to remain the same in the ideal case if there is no change in regulations like social distancing. It varies when there is a change in social distancing regulations or with variation in higher-order population mobility as in India’s case. Due to the implementation of a nationwide lockdown, the country has seen a labour migration from states to states. It was a huge migration and affected the social distancing parameters. Therefore, implementing lockdown and then unlocking resulted in a variation in **R_0_value**. The following equation shows the time and then dependency of **R_0_value**:

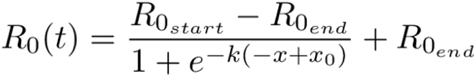

- **R_0start_** and **R_0end_** are the first and the last day values of **R_0_**
- **x_0_** is the value of x at the inflection point (the day recording steepest decline in the value of **R_0_**)
- **k** allows us to vary how quickly **R_0_** declines

## Resource- and Age-Dependent Fatality Rates

The fatality rate αdepends on a number of parameters such as health infrastructures in a state. For this study, we only consider the dependency on health resources and the average age of the infected population.

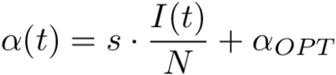

Here, *s* is some arbitrary but fixed (that means we choose it freely once for a model and then it stays constant over time) scaling factor that controls how big of an influence the proportion of infected should have; α_OPT_ is the optimal fatality rate. For example, if s = 1 and half the population is infected on one day, then s ⋅ I(t) / N = 1/2, so the fatality rate α(t) on that day is 50% + α _OPT_. Or maybe most people barely have any symptoms and thus many people being infected does not clog the hospitals. Then a scaling factor of 0.1 might be appropriate (in the same scenario, the fatality rate would only be 5% + α_OPT_).

## Triage and Limited Resources (Extended Model)

Due to limited resources available, Governments are adapting to triage especially in India and other highly impacted places. Doctors are differentiating between critical and normal patients in the wake of limited ICU Beds. This needs to be incorporated into the SEIR model for better prediction. If a country with B, ICU beds suitable to treat critical Covid-19 patients. If the amount C i.e. the number of critical patients more than B. This means that all the critical patients being admitted after filling up B ICU beds cannot be treated properly and thus might end up dead due to shortages. Thus we expanded our transition: from C, there are two populations we have to look at: max(0, C-B) people die because of shortages, and the rest get treated like we discussed above. Well, if C<B (enough beds), then C people get treated. If C>B (not enough beds), then B people get treated. That means that the remaining amount of people getting treatment is min(B, C) (again if C<B, then min(B, C) = C people get treatment; if C>B, then min(B,C) = B people get treatment; the math checks out).

Revising the model, with deaths happening immediately for all those above the number of beds available (one could change this to taking several days, dying with a probability of 75%, etc.). Instead of one(R_0_(t)), two time-dependent variables: R_0_(t) and Beds(t) are integrated into the model. For R_0_(t), the older logistic function will be used. While for Beds(t), the idea is that, as the virus spreads, countries react and start building hospitals, freeing up beds, etc. Thus, the number of beds available increases over time. Where Beds_0_ is the total number of ICU beds available and s is some scaling factor. In this formula, the number of beds increases by s times the initial number of beds per day (e.g. if s = 0.01, then on day t = 100, Beds(t) = 2 ⋅ Beds_0_)

**δ** and **γ** are fixed to δ = 1/9 and γ = 1/3 estimates based on acquired from [17]. Concerning **β(t)**, we’re calculating beta through R_0_(t) and γ, so there’s no need to find any separate parameters for the beta. The beds scaling factor **s** can be fitted; Admittedly, it does not play a big role in the outcome as, until now, the number of people not receiving treatment due to shortages was small compared to the total amount of deaths.

We collected two estimates for the probabilities **p(I→C)** and **p(C→D)**, split up by age group. This helped us calculate the probabilities, weighted by the proportion of the population per age group.

## Model Fitting

We needed a simple and flexible approach to modelling the data and curve fitting. It should allow the value of parameters to be constrained by an algebraic expression or vary over a range. Lmfit provides a high-level interface to non-linear optimization and curve-fitting problems for Python [10]. It seemed the best fit for the extended SEIR model. Data was integrated into and the known parameters’ boundary conditions were with initial guess value. To validate the use of the LMfit module in the above model, data of newly infected cases per day were compared to check if it was the best fit. And the model was perfectly valid.

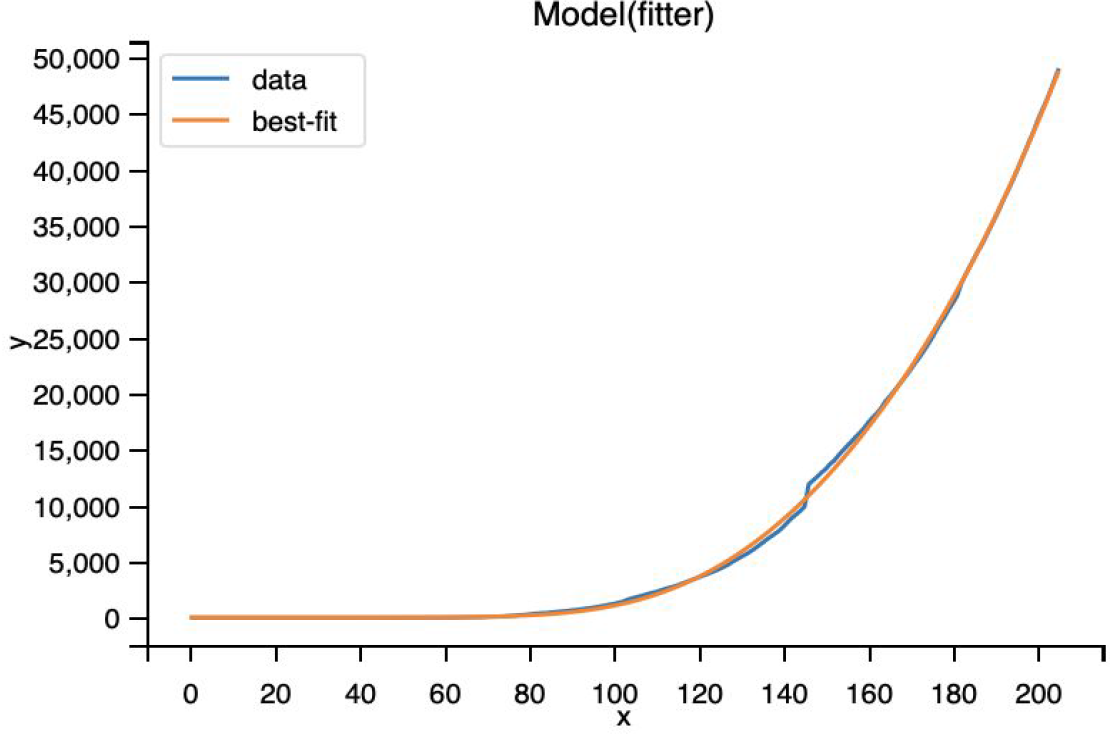

The best suitable parametric values for the extended SEIR model based on the analysed data are:

- R_0_ start: 3.041579778475209
- R_0_ end: 1.1418238404842034
- k: 0.05030810509474458
- p(C –> D): 0.7999984426717011
- p(I –> C): 0.0998597120800873
- s: 0.003
- x_0_: 77.38727582104808

We applied the LM-fit algorithm for parameter estimation in python and the code is available at https://github.com/brahmatheja123/Extended-SEIR-Model-.

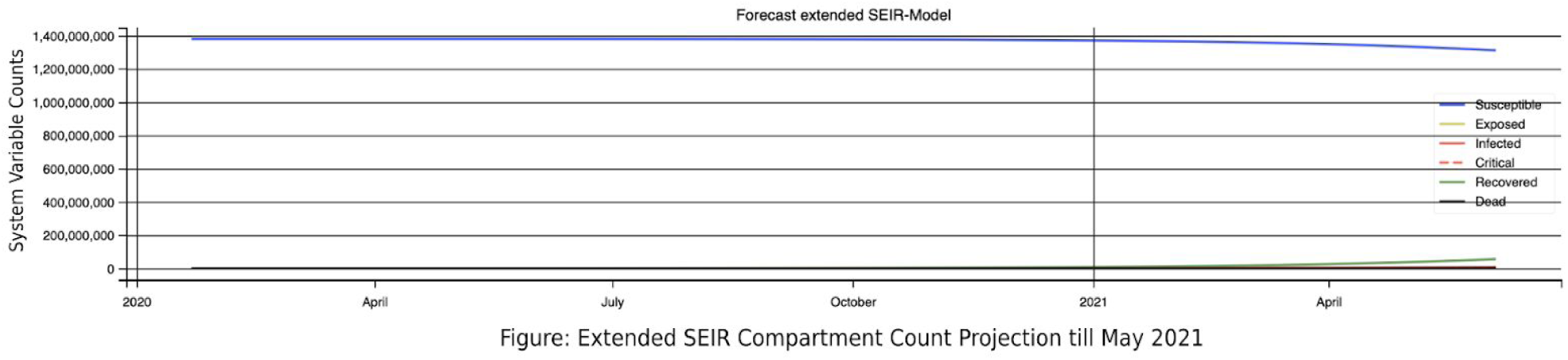

## Results

**Figure 1:**
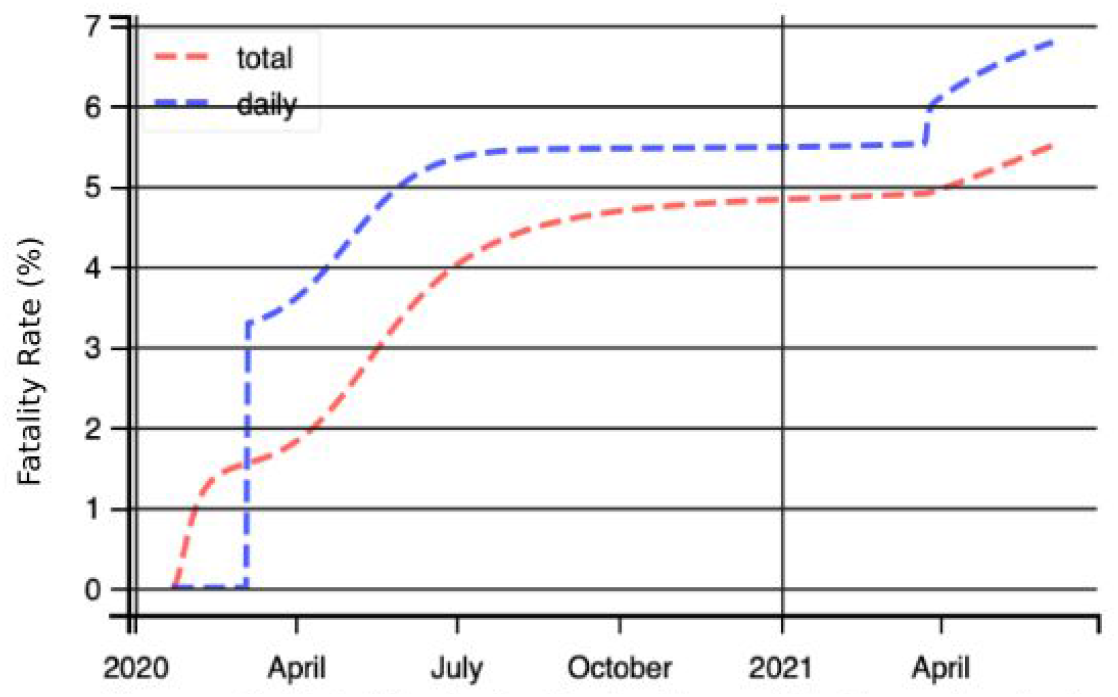
Fatality rate projection with time for India

The forecast for fatality rate is expected to be constant in the near future, proper handling of resources and instant availability of beds to the needed might keep the fatality rate at bay. Currently, the fatality rate is at 5.4%.

**Figure 2:**
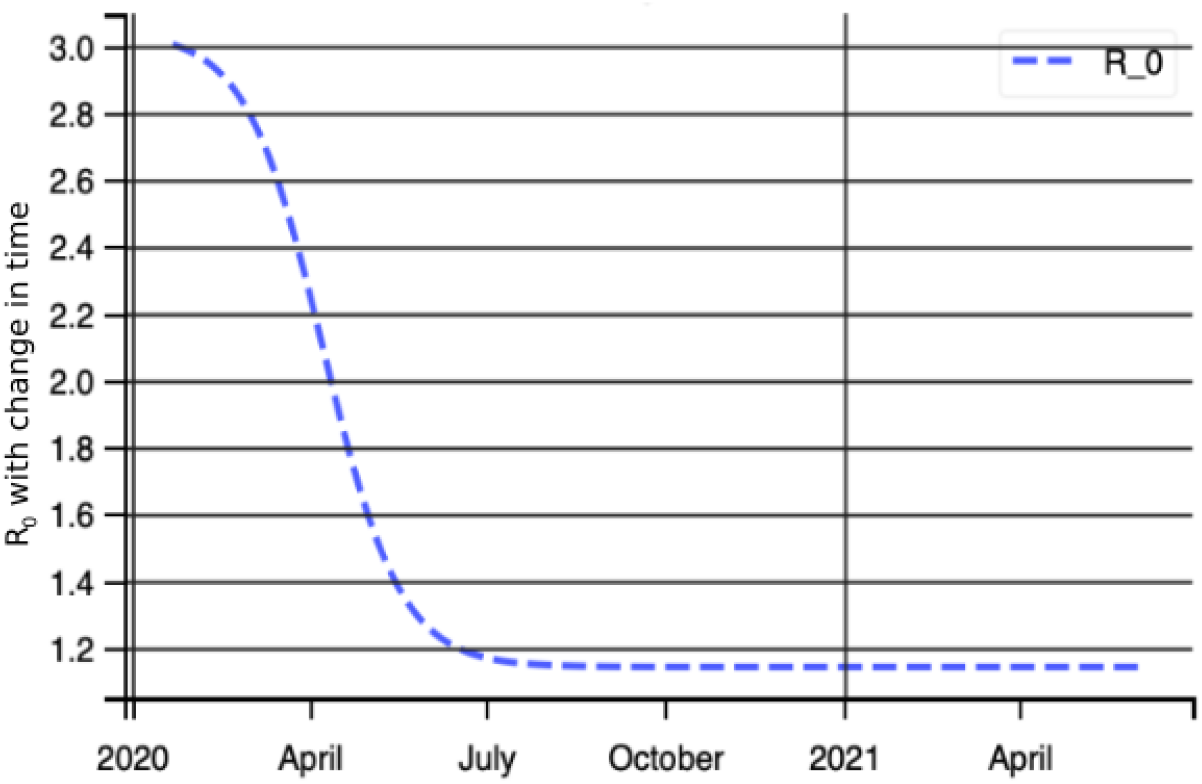
Year long projection of COVID-19 cases in India

Value of R_0_ had a steady decline since the start of complete lockdown in India and seems to pursue a similar value since the gradual opening of lockdown too. The forecast towards coping with this disease. But actual values may hinder the actual prediction in the near future if safety norms are not followed properly. Currently, the R_0_ value is at 1.17 for the overall country. This value is very high at places with high population density (like Delhi, Mumbai etc.) **citation for R_0 value of cities

Due to the limited availability of resources, they will exhaust themselves after a certain time. As we can see a gradual spike in Deaths per day from November onwards, with the daily count being at around 5000.

## Conclusion

Through study, we are able to construct a general understanding of transmission dynamics and what numbers we could expect in future considering the present scenarios. If the above forecast is right, India has not been through its worst yet. Though a decrease in R_0_ value, it does not mean COVID epidemic is under control. The percentage of infected patients being admitted into ICU for critical care is around 9.986%, while the chances of recovery of critical patients being admitted to the ICU seem to be slim at 79.9% of the admitted being dead. They need to prepare for more medical equipment, increasing ICU beds requirement and better physical distancing norms to keep the Deaths per day at bay. The goal was to forecast and prepare accordingly, at the same time understanding the seriousness of the situation to estimate and plan for hospital equipment and protective gear.

## Data Availability

OPEN SOURCE DATA ON COVID-19

## References

1. W.H. O, Statement on the second meeting of the International Health Regulations (2005) Emergency Committee regarding the outbreak of novel coronavirus (2019-nCoV). 2020.

2. Harapan H, Itoh N, Yufika A, et al. Coronavirus disease 2019 (COVID-19): A literature review. J Infect Public Health. 2020;13(5):667–673. doi:10.1016/j.jiph.2020.03.019

3. R. Singh et. al., Age-structured impact of social distancing on the COVID-19 epidemic in India, arXiv preprint 2020: arXiv:2003.12055v1 [q-bio.PE]

4. R. Ranjan, Predictions for COVID-19 outbreak in India using Epidemiological models, medRxiv 2020:https://doi.org/10.1101/2020.04.02.20051466

5. He, S., Peng, Y. & Sun, K. SEIR modelling of the COVID-19 and its dynamics. Nonlinear Dyn (2020). https://doi.org/10.1007/s11071-020-05743-y

6. Bouchnita, Anass, and Assam Jebrane. “A hybrid multi-scale model of COVID-19 transmission dynamics to assess the potential of non-pharmaceutical interventions.” Chaos, Solitons, and Fractals vol. 138 (2020): 109941. doi:10.1016/j.chaos.2020.109941

7. Killeen, Gerry F, and Samson S Kiware. “Why lockdown? Why national unity? Why global solidarity? Simplified arithmetic tools for decision-makers, health professionals, journalists and the general public to explore containment options for the 2019 novel coronavirus.” Infectious Disease Modelling vol. 5 442–458. 3 Jul. 2020, doi:10.1016/j.idm.2020.06.006

8. Coronavirus Outbreak in India, https://www.covid19india.org/

9. Floods in the time of coronavirus: Doubly whammy as Assam battles deluge, pandemic; Times Now Digital Updated Jul 16, 2020

10. Newville, Matthew; Stensitzki, Till; Allen, Daniel B.; Ingargiola, Antonino. “LMFIT: Non-Linear Least-Square Minimization and Curve-Fitting for Python”. https://doi.org/10.5281/zenodo.11813

11. Hou C, Chen J, Zhou Y, et al. The effectiveness of quarantine of Wuhan city against the Corona Virus Disease 2019 (COVID-19): A well-mixed SEIR model analysis. J Med Virol. 2020;92(7):841–848. doi:10.1002/jmv.25827

12. Adi-Kusumo F. The Dynamics of a SEIR-SIRC Antigenic Drift Influenza Model. Bull Math Biol. 2017;79(6):1412–1425. doi:10.1007/s11538-017-0290-5

13. Huo HF, Yang Q, Xiang H. Dynamics of an edge-based SEIR model for sexually transmitted diseases. Math Biosci Eng. 2019;17(1):669–699. doi:10.3934/mbe.2020035

14. Reno C, Lenzi J, Navarra A, et al. Forecasting COVID-19-Associated Hospitalizations under Different Levels of Social Distancing in Lombardy and Emilia-Romagna, Northern Italy: Results from an Extended SEIR Compartmental Model. J Clin Med. 2020;9(5):1492. Published 2020 May 15. doi:10.3390/jcm9051492

15. Rocklöv J, Sjödin H, Wilder-Smith A. COVID-19 outbreak on the Diamond Princess cruise ship: estimating the epidemic potential and effectiveness of public health countermeasures. J Travel Med. 2020;27(3):taaa030. doi:10.1093/jtm/taaa030

16. Peirlinck M, Linka K, Sahli Costabal F, Kuhl E. Outbreak dynamics of COVID-19 in China and the United States [published online ahead of print, 2020 Apr 27]. Biomech Model Mechanobiol. 2020;1–15. doi:10.1007/s10237-020-01332-5

17. Ministry of Health and Family Welfare. Government of India. MoHFW | Homemohfw.gov.in

